# TNFR Pathway-Related Proteins and Recurrent Coronary Artery Disease Events

**DOI:** 10.1101/2025.08.03.25332895

**Authors:** Jiwoo Lee, Tiffany R. Bellomo, Jennifer L. Halford, So Mi Jemma Cho, Satoshi Koyama, Jacqueline Dron, Sara Haidermota, Yunfeng Ruan, Sarah Urbut, Buu Truong, Aniruddh Patel, Pradeep Natarajan

**Author notes:** Authors contributed equally to this work (Jiwoo Lee, Tiffany R. Bellomo). **Corresponding Author:** Pradeep Natarajan, MD, MMSc, 185 Cambridge Street, CPZN 3.184, Boston, MA 02114.

## Abstract

Despite optimization with lifestyle modifications and medications, complications of coronary artery disease (CAD) remain the leading cause of adult mortality worldwide. This study aimed to identify proteins and pathways linked to recurrent CAD events to better understand residual risk. We used data from 1,009 participants in the UK Biobank (UKB) with baseline Olink plasma proteomic measures and CAD. Among 1,463 proteins tested, 102 proteins were independently associated with recurrent CAD events. Molecular functions were significantly enriched for tumor necrosis factor receptor (TNFR) activity by 100-fold (p-value = 6.37×10^-10^). Of the 16 proteins related to TNF annotated by Gene Ontology, TNF-alpha, TNFR1, and TNFR2 were all significantly associated with increased risk of recurrent CAD events. While TNFR1 and TNFR2 were initially thought to have opposing roles in cardiac remodeling post-MI, this study highlights the complex interaction between these pathways and the need to identify specific inflammation-related targets to therapeutic strategies.

## INTRODUCTION

Despite advances in prevention and treatment options, cardiovascular disease driven by coronary artery disease (CAD) has emerged as the leading cause of mortality and morbidity in the world in the 21^st^ century.^1^ Clinical practice guidelines continue to emphasize primary and secondary prevention of CAD through targeting modifiable risk factors, with an emphasis on lipid modification, blood pressure lowering, and glucose control.^2-4^ Despite optimization of these factors with lifestyle modifications and medications, the residual risk of recurrent CAD remains alarmingly high, with incidence dramatically increasing over extended follow-up periods.^5-7^ Recurrent CAD risk prediction models have demonstrated only a modest added benefit from inclusion of conventional biomarkers including N-terminal pro-brain natriuretic peptide (NT-proBNP),^8^ C-reactive protein (CRP),^9^ and troponin T,^10^ highlighting an area of unmet need.

The ongoing search for novel biomarkers that enhance risk stratification and mechanistic clarity has begun to increasingly leverage novel multi-plexed scalable platforms. Proteomics platforms enable broad-based scans to identify disease-relevant biomarkers. For example, utilization of these powerful technological approaches has improved predictive power for primary CAD,^11,12^ but there have only been limited assessments among those with CAD already manifested. A recent study modeling recurrent CAD events found that comprehensive clinical, lifestyle, sociodemographic, and genetic risk factors together accounted for less than 20% of the risk for CAD recurrence, suggesting the presence of yet unknown risk factors.^13^ The progression of CAD to recurrence involves numerous changes, including oxidative stress, endothelial cell dysfunction, and both pro-inflammatory and anti-inflammatory mechanisms.^14^ Many of these cellular changes may manifest in circulating plasma proteins.^15^

In this study, we investigated the plasma proteomic profiles of 1,009 participants from the UK Biobank (UKB) with CAD to identify associations with recurrent CAD events. We characterized the proteomic profiles in subgroups and enriched pathways associated with recurrent CAD events to better understand drivers and mechanisms underlying recurrent CAD events.

## RESULTS

### Baseline Characteristics

We included 1,009 participants from the UKB who had CAD at enrollment (Supplemental Figure 1). Participants had a mean age of 62.51 years (SD 5.94) at enrollment; 183 (18.14%) were female, and 656 (65.01%) had recurrent CAD events over 11.40 [interquartile range 8.00-14.69] years follow-up (Table 1). Those with versus without recurrent CAD events in follow-up had a higher rate of statin prescriptions (94.36% vs 89.52%, p-value = 0.007), a greater prevalence of diabetes (94.36% vs 89.52%, p-value = 0.007), and lower HDL cholesterol levels (44.01 mg/dl vs 45.84 mg/dl, p-value = 0.014). The remaining features stratified by recurrent CAD status were comparable within each cohort. Of the 52,705 UKB participants with 1,463 protein measurements, 42 proteins were imputed due to missing values.

**Table 1.**
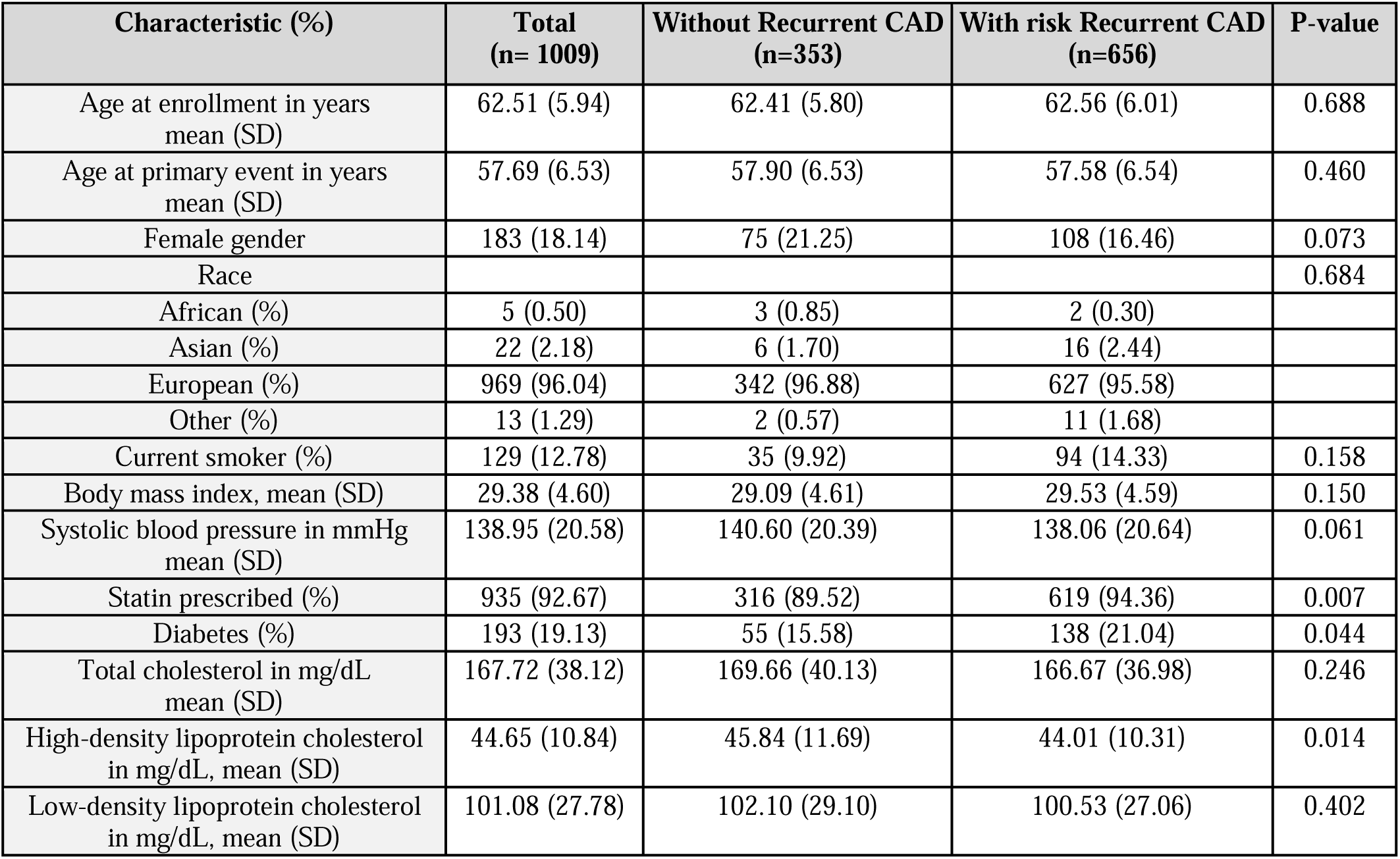
Demographics of the UK Biobank cohort by recurrent CAD status.

### Enrichment Analysis highlights the role of TNF-related pathways in Recurrent CAD

Among the 1,463 proteins tested, 482 proteins at nominal significance (p<0.05) and 102 proteins at experiment wide significance (Bonferroni p-value < 3.42×10^-5^) were associated with recurrent CAD events after adjusting for standard cardiovascular risk factors (Figure 1A, Supplemental Table 2). Of the 102 Bonferroni-corrected proteins associated with recurrent CAD events, 29 (28.43%) were classified as cardiometabolic proteins by Olink annotations (Supplemental Figure 2). Enrichment analysis of significant proteins using Gene Ontology (GO) terms revealed that molecular functions were significantly enriched for tumor necrosis factor receptor (TNFR) activity by 100-fold (p-value = 6.37×10^-10^) and death receptor activity by 100-fold (p-value = 9.30×10^-14^) (Figure 1B, Supplemental Figure 3). Annotation with GO terms for biological processes showed significant enrichment for the tumor necrosis factor-mediated signaling pathway (p-value = 2.04×10^-7^) (Supplemental Figure 4). GO terms for cellular compartments showed significant proteins were enriched in the endolysosome lumen (p-value = 2. 36×10-4) (Supplemental Figure 5).

**Figure 1.**
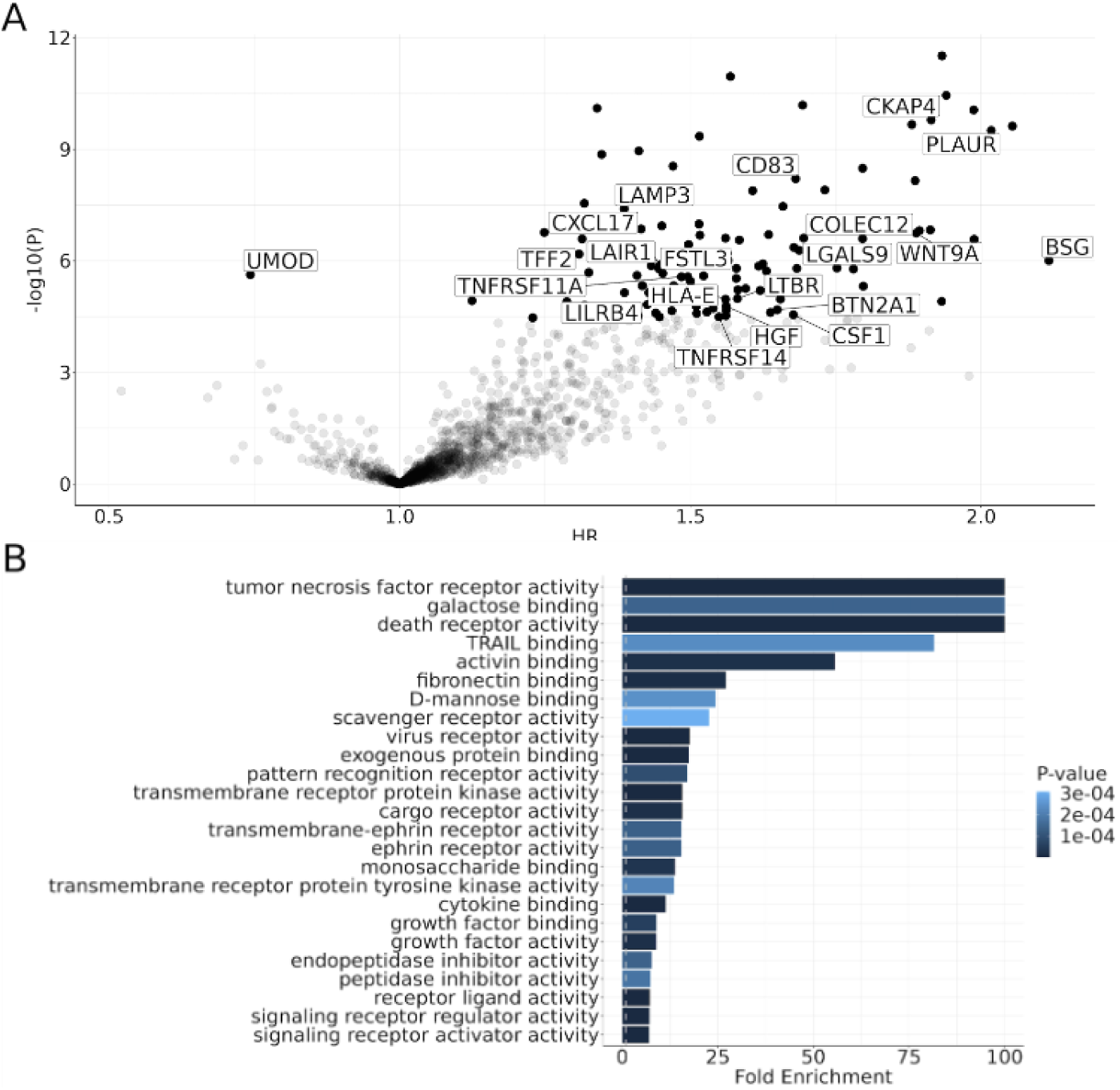
Risk of recurrent CAD events associated with measured proteins in the UKB annotated by Gene Ontology. (A) Volcano plot of hazard ratios and corresponding 95% confidence intervals are based on Cox proportional hazards regression models with covariates of age at UKB enrollment, age at first CAD event, sex, smoking status, diabetes diagnosis, body mass index, systolic blood pressure, LDL cholesterol, HDL cholesterol, statin prescription, and the first 10 principal components of genetic ancestry. All p-values were Bonferroni corrected for 1,463 measured proteins. DAVID annotation was utilized to provide Gene Ontology enrichment analysis of significant proteins by the top (B) molecular function.

To further investigate the role of TNF and related proteins annotated by signaling pathways, proteins were classified based on their primary involvement with TNFR1, TNFR2, both, or neither of these pathways and plotted alongside pathway annotations using the DAVID annotation tool (Figure 2, Supplementary Figure 5). GO term biological process identified 13 significantly enriched proteins in the apoptotic pathway, 16 significantly enriched proteins in the TNF-mediated signaling pathway, and 8 significantly enriched proteins in the canonical NF-κB pathway associated with increased risk of recurrent CAD events (Figure 2A, 2B, 2C). There was a similar finding when using KEGG pathway annotations, where TNF-alpha and TNF-related proteins were further implicated in cytokine-cytokine receptor interactions that occur in both apoptosis and canonical NF-κB signaling (Figure 2B). Within these clusters, TNF-alpha exhibited a risk estimate of 1.36 (95% CI 1.17-1.57, p-value = 6.38×10^-5^) for recurrent CAD events. TNFR1 (*TNFRSF1A*) had an HR of 1.73 (95% CI 1.43-2.09, p-value = 1.23×10^-8^) for developing recurrent CAD. Enriched proteins related to TNFR1, including RELT (HR 1.99, 95% CI 1.61-2.45, p-value = 8.71×10^-11^), TNFRSF21 (HR 1.64, 95% CI 1.30-1.44, p-value = 2.43×10^-5^), and TNFRSF11A (HR 1.48, 95% CI 1.26-1.75, p-value = 2.66×10^-6^) were also strongly associated with recurrent CAD events. TNFR2 (*TNFRSF1B*) had an HR of 1.27 (95% CI 1.13-1.44, p-value = 9.15×10^-5^) for developing recurrent CAD. Enriched proteins related to TNFR2, including TNFRSF14 (HR 1.55, 95% CI 1.26-1.90, p-value = 3.19×10^-5^) and TNFRSF9 (HR 1.27, 95% CI 1.20-1.60, p-value = 7.12×10^-6^) had a modest association with recurrent CAD events. Enriched proteins involved in both TNFR1 and TNFR2 pathways, including LTBR (HR 1.58, 95% CI 1.29-1.94, p-value = 1.03×10^-5^) and TNFRSF19 (HR 1.58, 95% CI 1.33-1.89, p-value = 2.74×10^-7^) were also associated with recurrent CAD events. A similar strength of association for all TNFR-related proteins was observed when median follow-up time of 11.40 years was limited to 3, 5, and 10 years (Supplemental Figure 6). A comprehensive list of annotated proteins with enriched UP-related terms is provided in Supplemental Figure 7 for sequencing features and Supplemental Figure 8 for biological processes.

**Figure 2.**
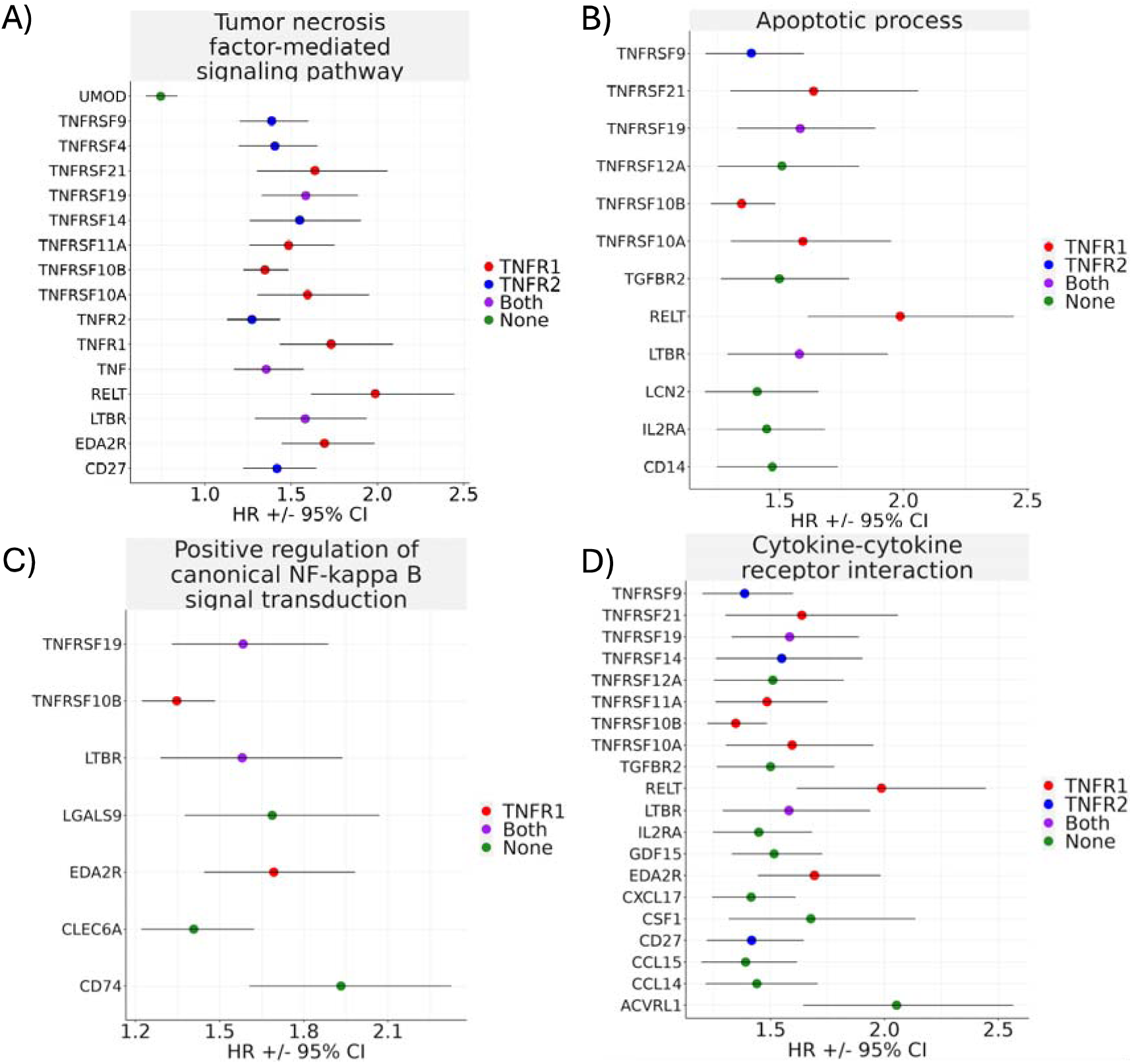
Proteins significantly associated with recurrent CAD events annotated using DAVID tools. Annotations were performed by (A) (B) (C) biological processes in the Gene Ontology database and (D) KEGG functional pathways. Hazard ratios and corresponding 95% confidence intervals are based on Cox proportional hazards regression models with covariates of age at UKB enrollment, age at first CAD event, sex, smoking status, diabetes diagnosis, body mass index, systolic blood pressure, LDL cholesterol, HDL cholesterol, statin prescription, and the first 10 principal components of genetic ancestry. Proteins were colored by involvement in TNFR1 as red, TNFR2 as blue, both pathways as purple, and neither of these two pathways in green.

### TNFR1 and TNFR2 Pathways Remain Associated with Recurrent CAD Across Subgroups

In exploratory analyses, we created survival plots for TNF-alpha and enriched TNF-related proteins, specifically those involved in TNFR1, TNFR2, or both pathways, that had the strongest association with recurrent CAD events. While in general, participants with elevated TNFR-related proteins had lower recurrence-free survival, confidence intervals were wide and the differences were not significant (log rank P>0.05) (Figure 3). We performed regression modeling of these significant TNF-related proteins in relation to recurrent CAD stratified by clinical subgroups to assess consistency of the observed associations across varying patient characteristics (Figure 4, Supplemental Table 3). There was a modestly higher risk of recurrent CAD events in younger patients compared to older for TNF-alpha (1.74 vs 1.25, p-interaction = 0.027) and TNFR2 (1.19 vs 1.64, p-interaction = 0.030), but not TNFR1 (1.66 vs. 1.80, p-interaction = 0.660). There was also a higher risk of recurrent CAD events in males compared to females for TNF-alpha (1.78 vs 1.17, p-interaction = 0.010) and TNFR2 (1.71 vs 1.07, p-interaction = 0.002), but not TNFR1 (1.79 vs 1.83, p-interaction = 0.590). Otherwise, the risk of recurrent CAD related to other TNF-related proteins remained similarly elevated among subgroups stratified by age, sex, race, and presence of diabetes. When comparing the prognostic value of TNF-related proteins to that of established cardiovascular risk factors, we found that the hazard ratios associated with TNF-related proteins were of comparable or greater magnitude, even after adjusting for all traditional risk factors (Supplemental Table 4). Among the conventional risk factors evaluated, statin use demonstrated the highest hazard ratio at 1.48 (95% CI: 1.05–2.09; p = 0.026). Notably, TNFR1 exhibited an even greater hazard ratio of 1.73 (95% CI: 1.43–2.09; p < 0.0001) following multivariable adjustment.

**Figure 3.**
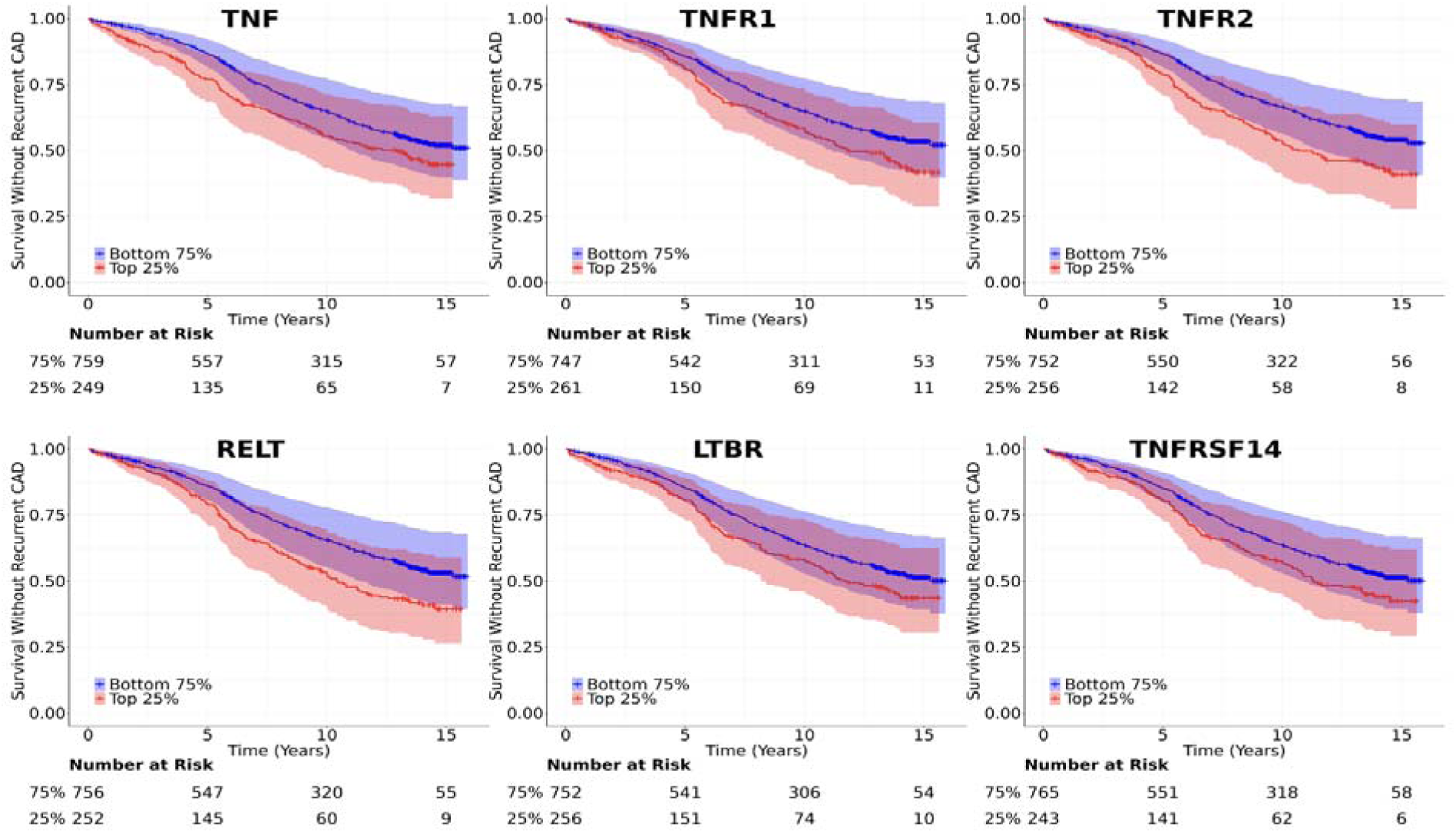
Kaplan–Meier curves showing recurrence-free survival for coronary artery disease (CAD) events during follow-up stratified by concentration of significantly enriched TNF-related proteins. Participants with protein concentrations in the highest quartile (Q4; red) were compared to those with lower concentrations (Q1–Q3; blue) using the log-rank test.

**Figure 4.**
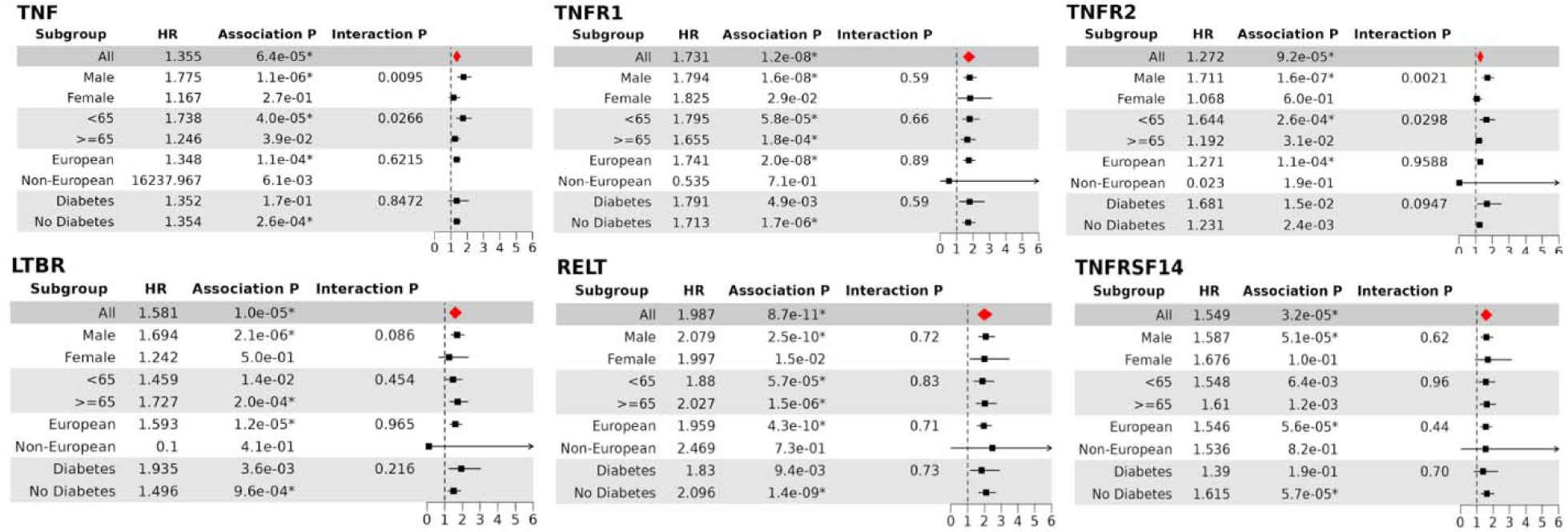
TNF-related proteins significantly associated with recurrent CAD events stratified by clinical subgroups. Hazard ratios and corresponding 95% confidence intervals are based on Cox proportional hazards regression models with covariates of age at UKB enrollment, age at first CAD event, sex, smoking status, diabetes diagnosis, body mass index, systolic blood pressure, LDL cholesterol, HDL cholesterol, statin prescription, and the first 10 principal components of genetic ancestry. Subgroup analyses were performed by splitting cohorts based on presence or absence of clinical grouping. An interaction p-value was calculated to identify significant interactions between clinical subgroups and proteins influencing recurrent CAD risk.

## DISCUSSION

In this study of 1,009 UKB participants with baseline CAD, we identified 102 proteins significantly associated with recurrent CAD events after adjustment for common cardiovascular risk factors. The identified proteins were significantly enriched for TNFR activity, cytokine-cytokine receptor interaction, and canonical NF-κB activation pathways. We demonstrate that the enriched TNFR proteins are positively associated with recurrent CAD events and span across both TNFR1 and TNFR2 pathways. These positive associations remained consistent across clinical subgroups of race and diabetes but suggest a potential effect modification related to age and sex for specific TNFR proteins.

Recurrent CAD event rates vary widely in the literature, depending on the duration of follow-up. At 3 years, Spironolactone trial reported event rates of 7.9% in the treatment group and 8.3% in the comparison group.^20^ By 5 years, the CARE trial demonstrated increasing recurrent event rates of 10.2% in the pravastatin group and 13.2% in the placebo group.^6^ A 2017 study using the UK Biobank identified 6,440 individuals with baseline CAD and of those, 3,733 experienced a recurrent event during the follow-up period at approximately 58%.^7^ Similarly, our analysis identified 656 participants (65.01%) with recurrent CAD events over a median follow-up of 11.40 years (IQR: 8.00–14.69). Restricting follow-up to 7, 5, or 3 years did not significantly alter the hazard ratios associated with TNFR-related proteins.

While the TNFR1 pathway has been associated with detrimental cardiac remodeling post-MI and the TNFR2 pathway with promoting cardiac recovery, this study revealed that elevated TNFR1 and TNFR2 correlate with an increased risk of recurrent CAD events. TNFR1 and TNFR2 can both be activated by TNF-alpha, released in large amounts from ischemia and hypoxia-activated cardiomyocytes following acute MI.^21^ TNFR1 activation is known promote apoptosis of certain cell types through the assembly of large signaling complexes, such as canonical activation, which collectively contribute to adverse cardiac remodeling.^22,23^ In contrast, TNFR2 is thought to enhance cardiac remodeling by activating cell survival pathways primarily though non-canonical NF-κB pathways, which mostly results in cell survival and proliferation.^24,25^ Recent research has uncovered overlap between TNFR1 and TNFR2 signaling, as both pathways share downstream signal proteins such LTBR and TNFRS19, which were also associated with recurrent CAD events in our study. These proteins activate the non-canonical NF-κB pathway that was enriched in this study,^26^ but the resulting effects depend on which of the 500 genes it transcriptionally regulates.^27^ Correspondingly, VSMCs can be signaled to survive, proliferate, and migrate to the areas of endothelial cell death causing increased intimal thickening and progression of atherosclerosis.^28,29^ Our study indicated that both TNRF1 and TNRF2 signaling are associated with increased risk of recurrent CAD events, underscoring the complex interaction between these two signaling pathways and the need to identify specific inflammation-related targets within these pathways to improve therapeutic strategies.

Cellular assays have demonstrated that TNFR2 signaling is dependent on the structure of TNFR2 itself. Vascular endothelial cells exhibit activation of NF-κB reporter genes when TNFR1 or TRAF2 binding sites of TNFR2 are expressed, but not when the constitutively active form of a TNFR2 domain Etk-SK is expressed alone.^30^ While certain forms of TNFR2, including those with TRAF binding sites, can activate the NF-κB reporter gene, this study did not show an association between TRAF2 and recurrent CAD events. This suggests that another subtype of TNFR2 may be responsible for triggering NF-κB activation. These findings emphasize the critical importance of elucidating TNFR2 structures and other associated structural proteins identified in this study to better understand its dual function in apoptotic mechanisms.

Emerging evidence highlights the clinical relevance of soluble TNF-related proteins in CAD, linking them to inflammation, cardiovascular events, and disease progression.^31^ In a study of humans with stable coronary heart disease, increased concentrations of both soluble TNFR1 and TNFR2 were associated with increased risks of cardiovascular events and mortality.^32^ While a cohort study adjusting for eGFR attenuated the association between TNFR1 and 2 with cardiovascular mortality,^33^ revascularization of cardiac lesions have also been shown to reduce serum concentrations of both TNF-alpha and serum levels of TNFR1, suggesting a functional relationship between soluble TNF-related proteins and inflammatory signaling.^34^ This functional relationship is further supported by studies of inflammatory diseases at both the cellular^35^ and patient levels.^36^ Our study identified associations not only with TNFR1 and TNFR2 but also with additional TNFR-related proteins, including TNFSF14. Soluble TNFSF14, that enters serum after proteolytic processing at the membrane, has previously been associated with repeat cardiovascular events in patients with stable CAD.^37^ Our study expands on this work by identifying an additional association between LTBR, a ligand of TNFSF14,^37^ and recurrent CAD events, highlighting the potential functional role of this pathway in CAD progression. Additionally, we demonstrate that the hazard ratios associated with the 16 TNF-related proteins were comparable to, or exceeded, those of traditional cardiovascular risk factors. These findings suggest that TNF-related proteins provide independent prognostic information beyond that captured by traditional risk factors.

Further work needed to identify which patient demographics are most likely to benefit from therapies targeting parts of the TNFR pathway. For example, males are known to have higher TNF-alpha levels compared to females with atherosclerotic disease.^38^ *In vitro* studies have also showed human neutrophils from males release more TNF-alpha than females when stimulated by the same amount concentrations of the stimulated by lipopolysaccharide.^39^ Our study similarly demonstrates that the association of TNF and recurrent CAD events is modified by sex, with males being at greater risk for recurrent CAD events and may represent a more responsive population to therapeutic agents. Additionally, we found TNF and TNFR2 had significant interaction terms in age groups, where elevated levels in younger individuals confer a higher risk for recurrent CAD events. This age-related susceptibility is supported by previous findings, such as the Northern Manhattan Stroke Study, which found that associations between TNF-related proteins and carotid stenosis were more prominent in relatively younger individuals.^40^ Similarly, mouse studies have shown that administering TNF-alpha treatment in young animals induces endothelial dysfunction, oxidative stress, and increased apoptosis in carotid and coronary arteries.^41^

Our study should be interpreted in the context of several limitations. First, majority of study participants were of European descent, a known limitation of the UKB, that compromises the generalizability of this study to diverse ancestries. We performed several subgroup analyses to assess for effect modification without significant differences in risk estimates. Second, plasma proteins of low abundance as well as cellular processes not reflected in plasma proteomics are not suitably assessed through the present technology. Third, this study did not account for medications that influence TNFR levels, as the UKB may not have comprehensively recorded all such medications. Fourth, we used the Olink technology from a single cohort. Nevertheless, our observations aligned with several experimental models assessing the role of TNFR-related proteins on atherosclerosis.

A comprehensive proteomic assessment of recurrent CAD events prioritized 102 proteins, with a notable enrichment of TNF-related proteins, associated with recurrent CAD events. Elevated TNFR1, TNFR2, and proteins associated with both pathways correlated with an increased risk of recurrent CAD events. While TNFR1 and TNFR2 were initially thought to have opposing roles in cardiac remodeling post-MI, this study provides evidence of the complex interaction between these signaling pathways and highlights the need to identify specific inflammation-related targets to improve therapeutic strategies.

## METHODS

### Study population

The UKB is a prospective cohort study with genetic and clinical data collected on approximately 500,000 individuals between the ages of 40 and 69 years at the time of recruitment.^16^ The data were linked to the National Health Service records from 2006-2022, permitting identification of prevalent clinical conditions and incident events. Analyses in the UKB were approved by the Northwest Multi-centre Research Ethics Committee (11/NW/0382) and secondary use scope (application #7089) by the Massachusetts General Hospital Institutional Review Board in accordance with the Declaration of Helsinki under IRB #2021P002228.

Based on physician diagnoses or procedural codes, 10,688 participants had CAD prior to UK Biobank enrollment (Supplemental Figure 1). We then excluded 322 participants with unavailable genotype information and 2,067 individuals with incomplete covariates measurements. An additional 7,270 individuals did not have proteomic profiling at baseline and were excluded. The final analytical cohort included 1,009 individuals.

### Outcome measure

Prevalent CAD was defined with previously utilized diagnoses codes for a CAD event identified in the HESIN master table that entails information on inpatient episodes of care, including diagnoses, admissions and discharge, operations, and procedures described in UKB (Supplemental Table 1). Specifically, diagnostic and procedural codes for a CAD event included myocardial infarction, percutaneous coronary intervention, coronary artery bypass graft, or the death register indicating myocardial infarction and related sequelae as either a primary or secondary cause of death in the UKB. The primary outcome of a recurrent CAD event in the UKB was defined as previously described,^13^ which briefly was the first CAD event that occurred after UKB recruitment irrespective of the number of CAD events prior to recruitment. The recurrent CAD event had to occur at least 28 days after the most recent CAD event prior to recruitment as previously described to differentiate independent incidences from combined attributions of diagnoses and procedures within a single episode or hospitalization.

### Variables

Other standard cardiovascular risk factors used in this analysis were selected based on previously described cardiovascular risk prediction models,^13^ including age at enrollment, age at first CAD event, sex, cigarette smoking, diabetes diagnosis, body mass index (BMI), systolic blood pressure, low-density lipoprotein (LDL) cholesterol, high-density lipoprotein (HDL) cholesterol, statin prescription, and the first 10 components of genetic ancestry. Both the age at UKB enrollment and the age at first CAD event were utilized in the analysis. Sex and racial background were both derived from fixed self-reported categories. Racial background included categories of African, Bangladeshi, British, Caribbean, Chinese, Indian, Irish, Pakistani, White and Asian, White and Black African, White and Black Caribbean, Other Asian, Other Black, Other White, Other mixed, or Other/unknown. Given small sample sizes of some groups, categories were consolidated into broader categories of African, Asian, European, and Other for analysis. Current cigarette smoking was defined as lifetime smoking of at least 100 cigarettes and currently without cessation. BMI was measured using Tanita BC-418MA body composition analyzer. Blood pressure was measured by the UKB staff at the time of enrollment. All laboratory values for total and HDL cholesterol were derived from a non-fasting sample collection assayed within 24 hours. Statin prescription was defined as a prescription written at the time of enrollment. Diabetes mellitus was defined as glycated A1c ≥6.5% or prior physician diagnosis.^17^

### Proteomics data processing

Proteomics data was collected from a random set of individuals enrolled in the UK Biobank Pharma Proteomics Project (UKB-PPP) as previously described.^18^ Briefly, non-fasting baseline plasma samples were collected from 52,749 UK Biobank participants using the Olink Explore 1536 platform with Olink Proximity Extension Assay technology. Exclusion criteria for individuals were high rate of missingness (>10% missing protein measurements) and those with excess relatedness (second-degree relatives or closer). The remaining missing values were imputed using the “miceforest” (Python package) with 10-fold cross-validation to impute the residual absent protein readings in the quality-controlled Olink proteomics dataset. While performing imputation, we evaluated factors that can potentially affect protein measurements such as protein batch, the study center, and first 10 principal components of genetic ancestry. Our evaluation indicated a negligible impact on protein levels arising from these factors, with the weakest Pearson correlation between protein levels and residuals being 0.93.

### Statistical testing

Cox proportional hazards regression models were run to assess the association between proteins measured and recurrent CAD events in a population of patients with CAD. Multivariate models were constructed, testing the association of each protein individually with recurrent CAD events. Consistent with prior studies, covariates included standard cardiovascular risk factors, and all p-values reported were two-sided. P-values were corrected using the corresponding Bonferroni p-value for multiple testing of proteins. Significant proteins were annotated with Gene Ontology (GO), Kyoto Encyclopedia of Genes and Genomes (KEGG), and UniProt (UP) associated terms and enrichment analysis was performed using the Database for Annotation, Visualization, and Integrated Discovery (DAVID) tool in the background of Olink proteins.^19^ The GO term enrichment analysis was performed with the DAVID online platform.

Kaplan Meier survival plots were constructed to evaluate time to recurrent CAD event in those with significant enriched proteins in the 4th quartile versus lower concentrations and compared using the log rank test. Cox proportional hazards regression models within demographic and comorbid subgroups were used to quantify the relationship between protein levels and time to recurrent CAD events. All statistical analyses were performed using R version 4.2.2 (R Foundation for Statistical Computing, Vienna, Austria. URL: https://www.R-project.org/).

## Data availability statement

Full individual-level data is available upon application to the UK Biobank.

## Acknowledgements

The authors would like to acknowledge the participants and staff of the UK Biobank. This work was conducted under UK Biobank application 7089. This work was supported by grants 1F32HL174327-01 (to T.R.B.), K99HL177340 (to S.M.J.C), K08HL168238 (to A.P.P.), R01HL127564 (to P.N.), from the National Heart, Lung, and Blood Institute; U01HG011719 (to A.P.P. and P.N.) from the National Human Genome Research Institute.

## Author contributions

J.L., T.R.B., J.L.H., and P.N. designed the study. J.L., T.R.B., J.L.H., S.M.J.C., S.K., J.D., S.U., B.T., and P.N. acquired, analyzed or interpreted data. J.L., T.R.B., and P.N. wrote the manuscript. S.M.J.C., S.K., J.D., Y.R., S.U., B.T., and A.P provided revisions.

J.L., T.R.B., S.H., and S.U. provided administrative, technical or material support.

## Competing Interests

P.N. reports research grants from Allelica, Amgen, Apple, Boston Scientific, Cleerly, Genentech / Roche, Ionis, Novartis, and Silence Therapeutics, personal fees from Allelica, Apple, AstraZeneca, Bain Capital, Blackstone Life Sciences, Bristol Myers Squibb, Creative Education Concepts, CRISPR Therapeutics, Eli Lilly & Co, Esperion Therapeutics, Foresite Capital, Foresite Labs, Genentech / Roche, GV, HeartFlow, Magnet Biomedicine, Merck, Novartis, Novo Nordisk, TenSixteen Bio, and Tourmaline Bio, equity in Bolt, Candela, Mercury, MyOme, Parameter Health, Preciseli, and TenSixteen Bio, royalties from Recora for intensive cardiac rehabilitation, and spousal employment at Vertex Pharmaceuticals, all unrelated to the present work. The remaining authors have nothing to disclose.

## Notes

### Author Declarations

Analyses in the UKB were approved by the Northwest Multi-centre Research Ethics Committee (11/NW/0382) and secondary use scope (application #7089) by the Massachusetts General Hospital Institutional Review Board in accordance with the Declaration of Helsinki under IRB #2021P002228.

