## Supplemental Figures 1-7 for "TNFR Pathway-Related Proteins and Recurrent Coronary Artery Disease Events"

**Enriched TNRF2 Pathway-Related Proteins Associated with Recurrent Coronary Artery Disease**

a) Program in Medical and Population Genetics and the Cardiovascular Disease Initiative,

Broad Institute of Harvard and MIT, Cambridge, MA

b) Division of Vascular and Endovascular Surgery, Massachusetts General Hospital, Boston, MA

c) Integrative Research Center for Cerebrovascular and Cardiovascular Diseases, Yonsei University College of Medicine, Seoul, Republic of Korea.

d) Center for Genomic Medicine, Department of Medicine, Massachusetts General Hospital, Boston, MA, USA

e) Division of Cardiology, Massachusetts General Hospital, Harvard Medical School, Boston, MA

**Table of Contents**

**Supplemental Tables:**

Supplemental Table 1: Definition of coronary artery disease in the UK Biobank.

Supplemental Table 2: Risk of recurrent coronary artery disease associated with measured proteins.

Supplemental Table 3: Risk of recurrent coronary artery disease associated with measured proteins for clinical subgroups.

Supplemental Table 4. Risk of recurrent coronary artery disease associated with traditional cardiovascular risk factors and TNFR-related proteins.

**Supplemental Figures:**

Supplemental Figure 1: Flowchart of inclusion and exclusion criteria in UK Biobank.

Supplemental Figure 2: Risk of recurrent CAD associated with measured proteins in UKB by functional category.

Supplemental Figure 3. Risk of recurrent CAD associated with measured proteins in UKB by molecular function annotated using Gene Ontology.

Supplemental Figure 4. Risk of recurrent CAD associated with measured proteins in UKB by biological processes annotated using Gene Ontology.

Supplemental Figure 5. Risk of recurrent CAD associated with measured proteins in UKB by cellular components using Gene Ontology annotations.

Supplemental Figure 6. Risk of recurrent CAD associated with measured proteins in UKB annotated by sequence feature in the UniProt database.

Supplemental Figure 7. Risk of recurrent CAD associated with measured proteins in UKB annotated by biological process in the UniProt database.

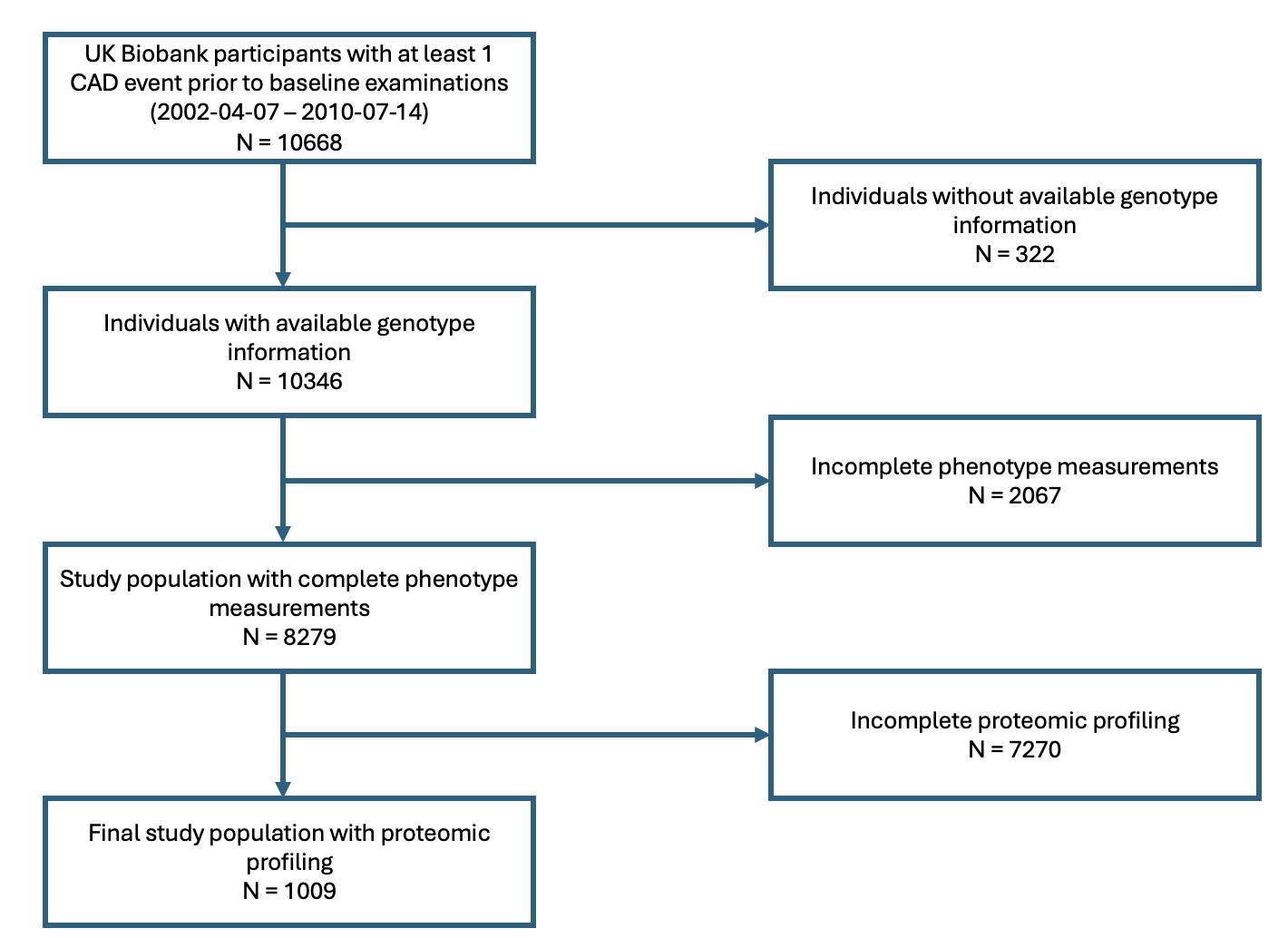

**Supplemental Figure 1.** Flowchart of inclusion and exclusion criteria in UK Biobank. Of the 502,679 participants enrolled in the UK Biobank, only a random sample of 52,705 underwent proteomic profiling. We focused our analysis only on individuals who already had an existing coronary artery disease event before their baseline enrollment exam, which resulted in a population of 1,256 participants.

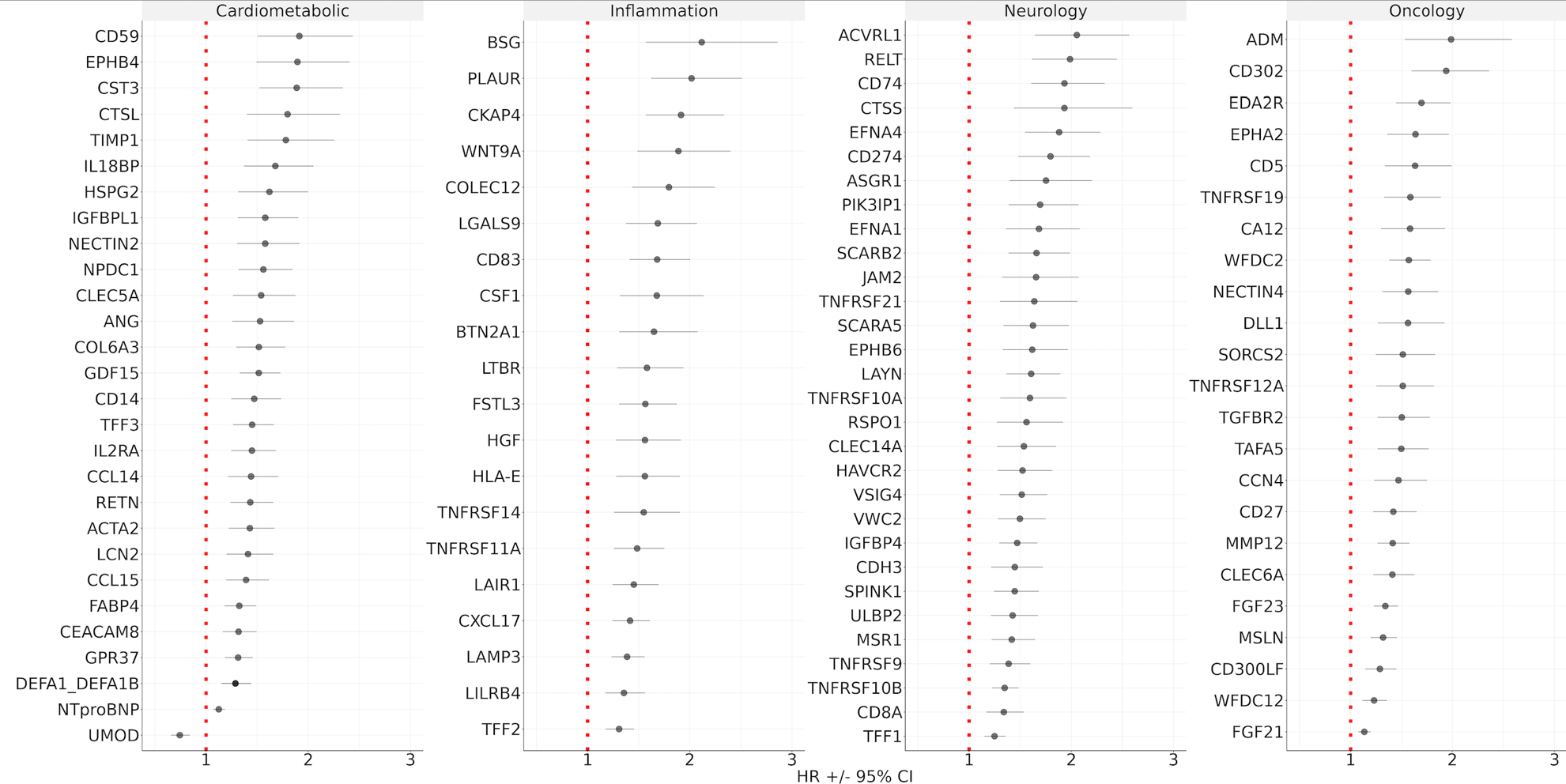

**Supplemental Figure 2.** Risk of recurrent CAD associated with measured proteins in UKB by functional category. Hazard ratios and corresponding 95% confidence intervals are based on Cox proportional hazards regression models with covariates of age at UKB enrollment, age at first CAD event, sex, smoking status, diabetes diagnosis, body mass index, systolic blood pressure, LDL cholesterol, HDL cholesterol, statin prescription, and the first 10 principal components of genetic ancestry.

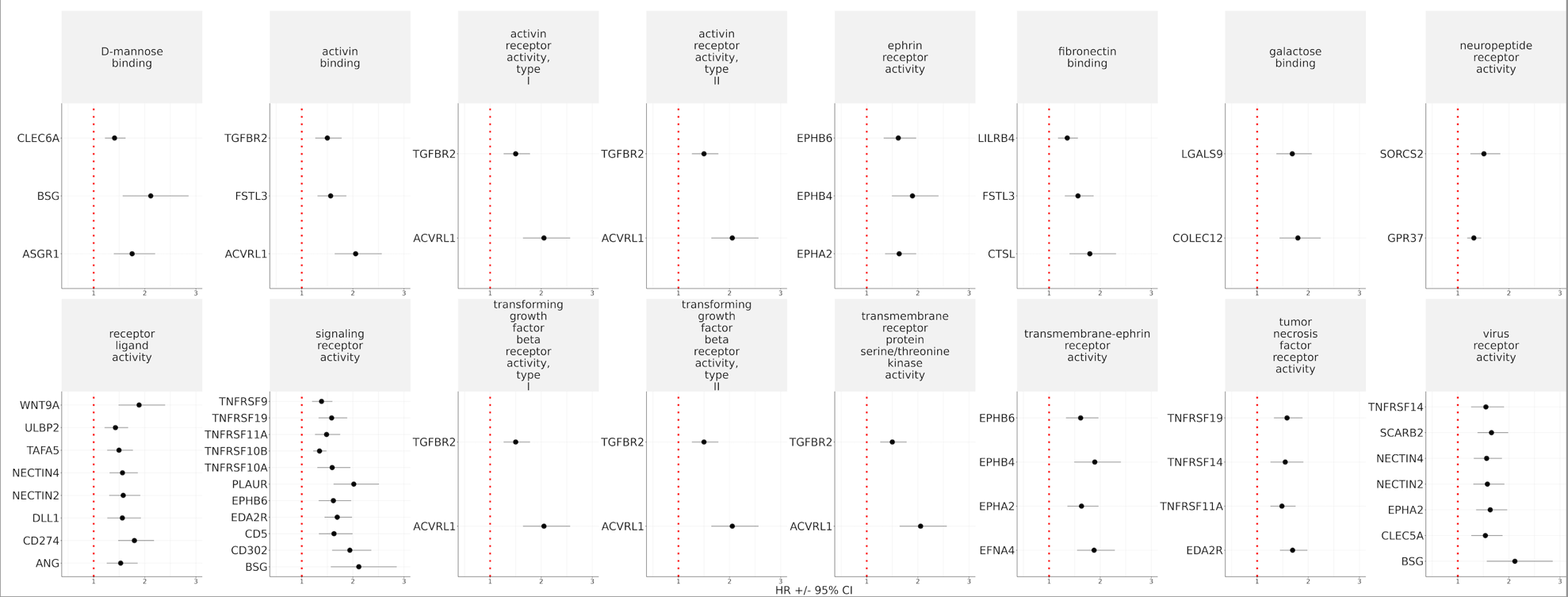

**Supplemental Figure 3.** Risk of recurrent CAD associated with measured proteins in UKB by molecular function annotated using Gene Ontology. Hazard ratios and corresponding 95% confidence intervals are based on Cox proportional hazards regression models with covariates of age at UKB enrollment, age at first CAD event, sex, smoking status, diabetes diagnosis, body mass index, systolic blood pressure, LDL cholesterol, HDL cholesterol, statin prescription, and the first 10 principal components of genetic ancestry.

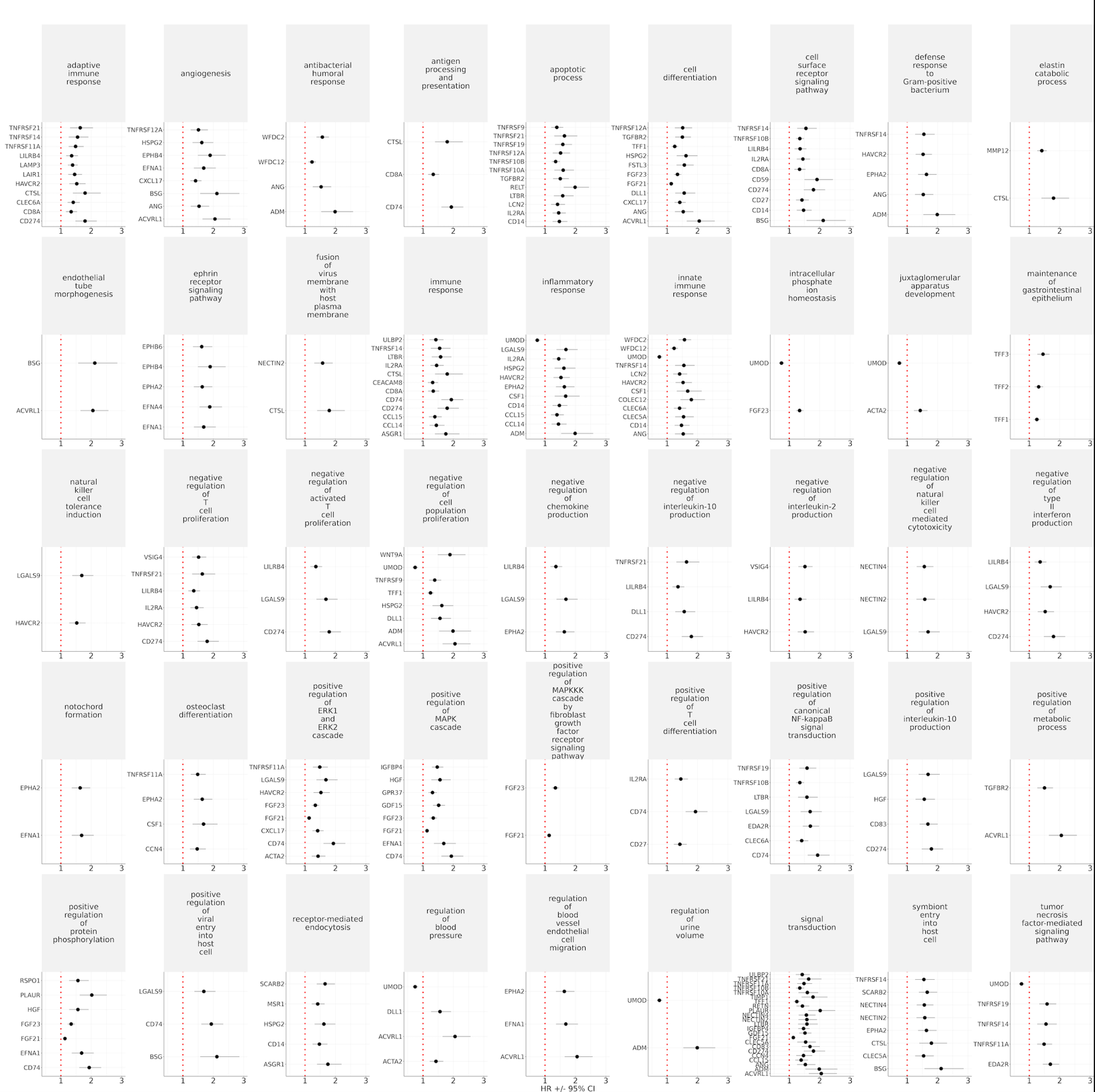

**Supplemental Figure 4.** Risk of recurrent CAD associated with measured proteins in UKB by biological processes annotated using Gene Ontology. Hazard ratios and corresponding 95% confidence intervals are based on Cox proportional hazards regression models with covariates of age at UKB enrollment, age at first CAD event, sex, smoking status, diabetes diagnosis, body mass index, systolic blood pressure, LDL cholesterol, HDL cholesterol, statin prescription, and the first 10 principal components of genetic ancestry.

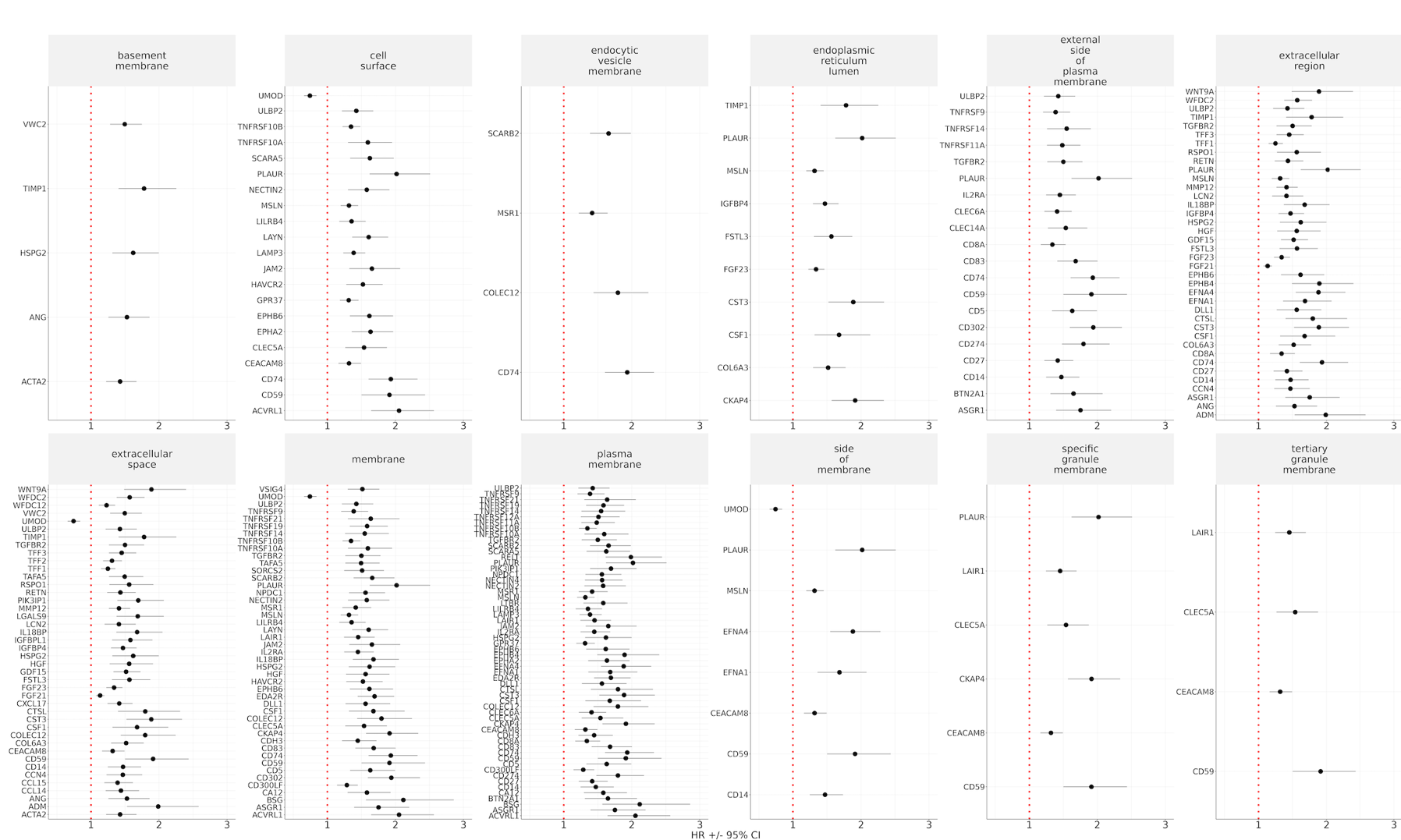

**Supplemental Figure 5.** Risk of recurrent CAD associated with measured proteins in UKB by cellular components using Gene Ontology annotations. Hazard ratios and corresponding 95% confidence intervals are based on Cox proportional hazards regression models with covariates of age at UKB enrollment, age at first CAD event, sex, smoking status, diabetes diagnosis, body mass index, systolic blood pressure, LDL cholesterol, HDL cholesterol, statin prescription, and the first 10 principal components of genetic ancestry.

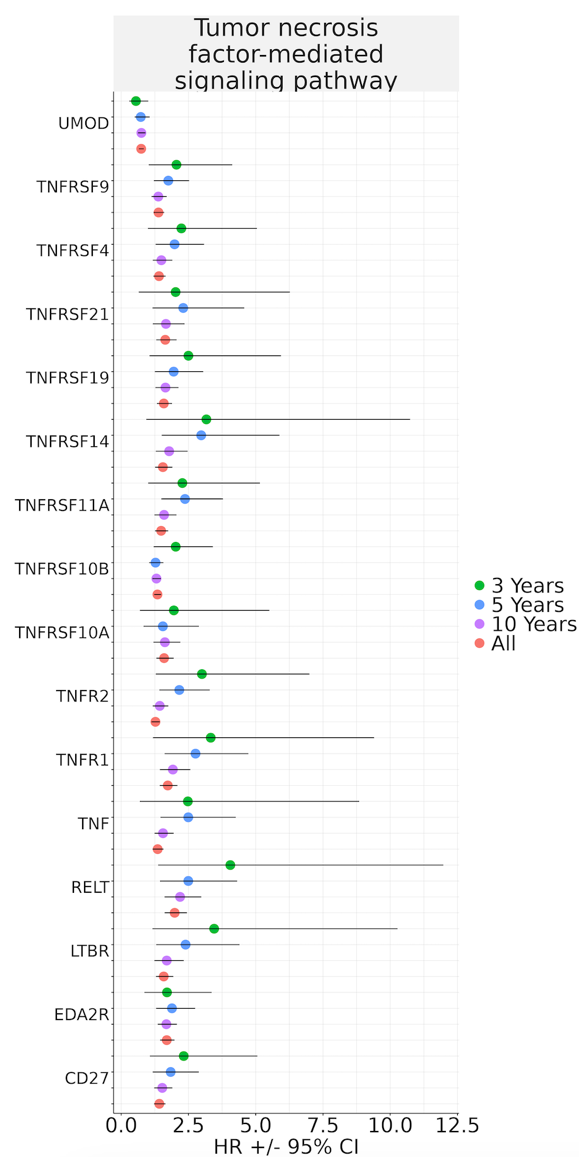

**Supplemental Figure 6.** TNFR-related proteins significantly associated with recurrent coronary artery disease (CAD) across varying follow-up durations. Proteins were colored by follow-up duration where green is 3 years, blue is 5 years, purple is 10 years, and red is the full length of follow up time. Hazard ratios and corresponding 95% confidence intervals are based on Cox proportional hazards regression models with covariates of age at UKB enrollment, age at first CAD event, sex, smoking status, diabetes diagnosis, body mass index, systolic blood pressure, LDL cholesterol, HDL cholesterol, statin prescription, and the first 10 principal components of genetic ancestry.

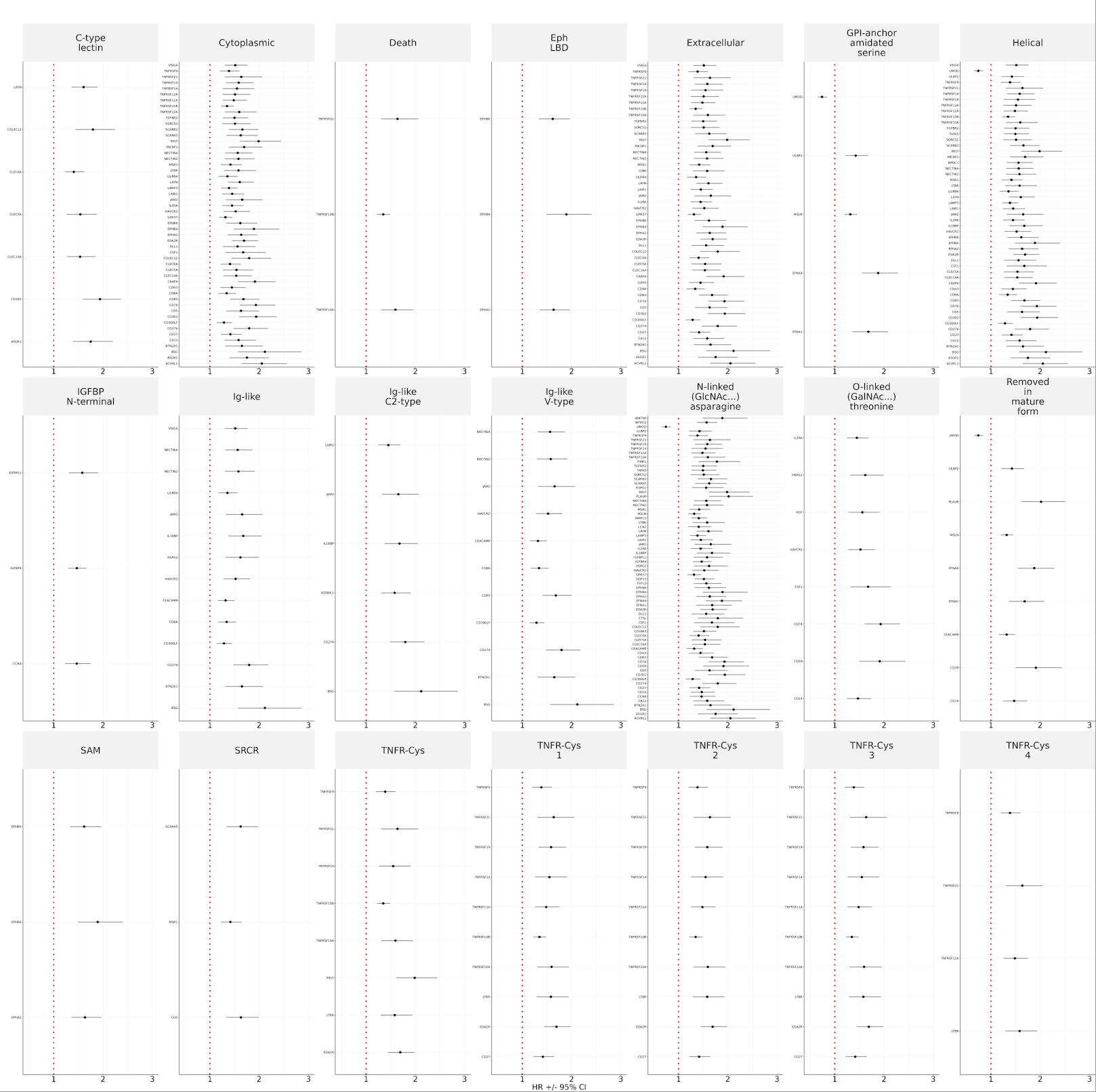

**Supplemental Figure 7**. Risk of recurrent CAD associated with measured proteins in UKB annotated by sequence feature (UP_SEQ_FEATURE) in the UniProt (UP) database. Hazard ratios and corresponding 95% confidence intervals are based on Cox proportional hazards regression models with covariates of age at UKB enrollment, age at first CAD event, sex, smoking status, diabetes diagnosis, body mass index, systolic blood pressure, LDL cholesterol, HDL cholesterol, statin prescription, and the first 10 principal components of genetic ancestry.

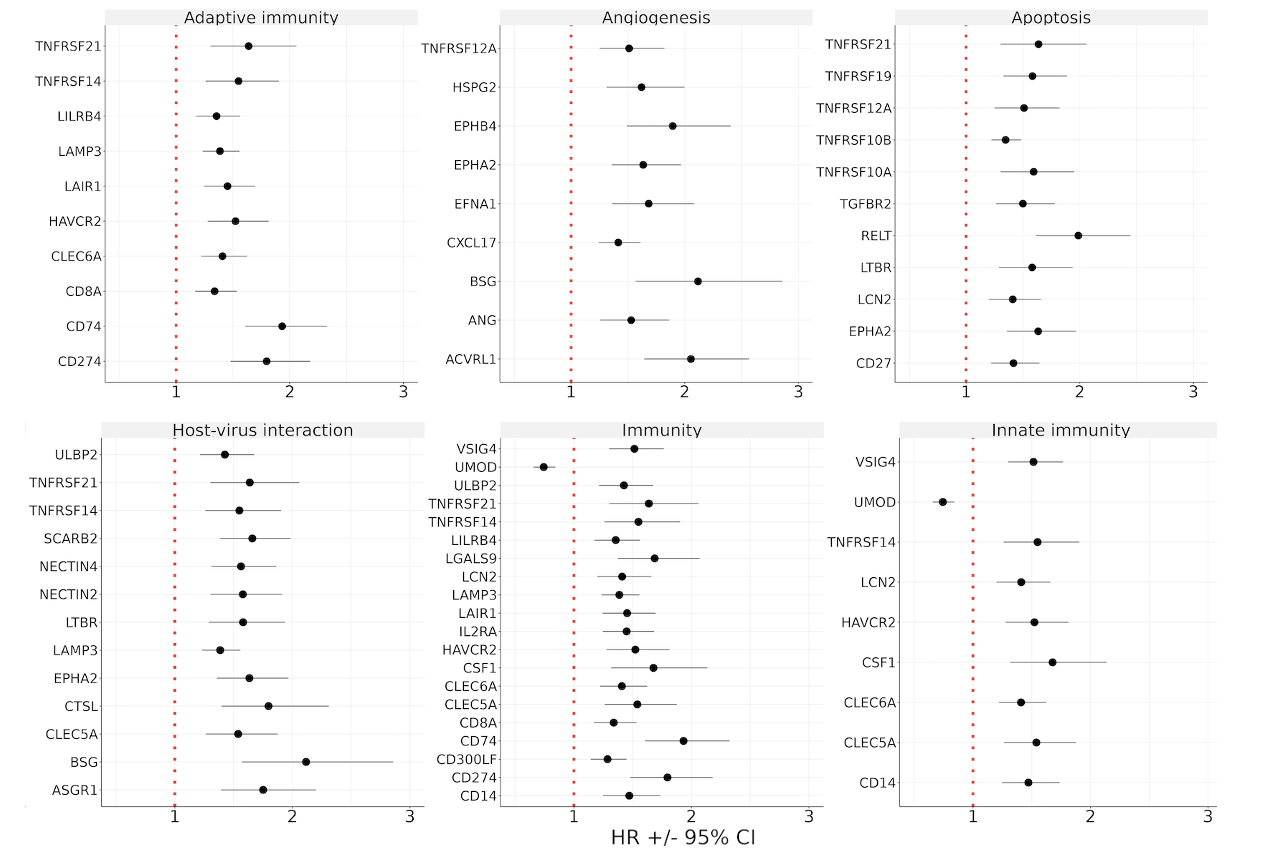

**Supplemental Figure 8**. Risk of recurrent CAD associated with measured proteins in UKB annotated by biological process (UP_KW_BIOLOGICAL_PROCESS) in the UniProt (UP) database. Hazard ratios and corresponding 95% confidence intervals are based on Cox proportional hazards regression models with covariates of age at UKB enrollment, age at first CAD event, sex, smoking status, diabetes diagnosis, body mass index, systolic blood pressure, LDL cholesterol, HDL cholesterol, statin prescription, and the first 10 principal components of genetic ancestry.
